# Clinical observation of high-flow nasal cannula with non-rebreather mask use on severe or critically ill COVID-19 diabetic patients

**DOI:** 10.1101/2021.10.13.21264946

**Authors:** AFM Tareq Bhuiyan, Sudipta Deb Nath, Md Jakir Hossain, Shuva Das, Moumita Das, Moinul Ahsan, Md. Iftekher-E-Alam Ziad, Fahmida Khatun Padma, Rana Dey, AKM Shamsul Alam, Farial Hoque Zehan, Ayan Saha

## Abstract

**Background and aims:** Prevalence of diabetes is a vital factor in COVID-19’s clinical prognosis. This study aimed to investigate and compare the efficacy of High-flow Nasal Cannula (HFNC) with/without non-rebreather mask (NRM) use on critical COVID-19 patients with/without diabetes.

**Methods:** For analysis and comparison, epidemiological, biochemical, and clinical data were collected from 240 HFNC (±NRM) treated severe and critical COVID-19 patients (diabetic = 136; non-diabetic = 104) admitted into ICUs of five hospitals in Chattogram, Bangladesh.

**Results:** 59.1% of patients with fever had diabetes (p=0.012). ICU stay was longer for diabetic patients (9.06±5.70) than non-diabetic patients (7.41±5.11) (p=0.020). Majority of the hypertensive patients were diabetic (68.3%; p<0.001). Majority of diabetic patients (70.4%; p<0.005) had elevated creatinine levels. Partial pressure of oxygen (mmHg) after HFNC (only) administration was significantly (p=0.031) higher in non-diabetic patients (69.30±23.56) than in diabetic patients (61.50±14.49). Diabetic (62.64±13.05) and non-diabetic patients (59.40±13.22) had almost similar partial pressure of oxygen (mmHg) from HFNC with NRM. Patients with elevated RBS required NRM with HFNC five times (AOR=5.1, 1.2-20.8) higher than others. Besides age, and hypertension were significantly associated with the HFNC+NRM treated diabetic patients. Factors those affected the HFNC only treated patients were fever and impaired glucose tolerance.

**Conclusions:** The results of this study imply that oxygen supply with HFNC and NRM may be beneficial for the elderly/hypertensive diabetic patients with COVID-19 associated AHRF; and that increased blood glucose level could be a determinant for the need of HFNC + NRM treatment.

**Highlights:**

1. Elderly diabetic patients required both HFNC and NRM to increase oxygen saturation.
2. Hypertension may be a factor for diabetic patients with COVID-19 requiring HFNC and NRM together.
3. ‘HFNC + NRM’-combination therapy might be needed when blood glucose levels rise.

## 1. Introduction

Coronavirus Disease 2019 (COVID-19) is a multisystem illness that majorly affects the respiratory tract; induced by a newer variant of the severe acute respiratory syndrome-related Coronavirus (SARS-CoV-2) [1]. The breakout of the then novel virus was detected in Wuhan, China by the end of December 2019 and in a very short span of time, the SARS-CoV-2 being highly contagious was able to rapidly spread between populations. The rather fast global spread of the disease and its severe clinical outcomes prompted the World Health Organization to proclaim it a pandemic on March 11, 2020 [2, 3].

During the outbreak, case-control studies on COVID-19 found that comorbid conditions like diabetes mellitus might predict COVID-19 advancement in patients [3]. Although the evidence is limited, recent research has suggested that diabetes and high blood sugar levels can operate as predictor variables in COVID-19-related disease burden; firstly, as because diabetic patients have a weakened immune system, they take longer to recover from viral infections, and secondly, since the virus may survive in a high-glucose condition. These denominators put people with diabetes in a susceptible position in terms of COVID-19 fatalities [4-6]. Furthermore, several COVID-19 related long-term sequelae have been reported in current research [7, 8], necessitating comprehensive inquiry and evaluation to confirm the evidence in depth.

Bangladesh ranks eighth among the world’s most populous countries, with almost 161 million people [9]. To date, more than 0.7 million infected cases have been reported in Bangladesh, while the lethalities have reached a count of more than 12 thousand (https://iedcr.gov.bd/). Diabetes, among other chronic disease states, seems to be on the upswing in Bangladesh at a rapid pace, with 8.4 million instances in adults, according to data from the International Diabetes Federation (IDF) [10]. A number of studies reported strong correlation between diabetes and COVID-19 [11, 12].

The high number of severe and critical COVID-19 cases has imposed an unprecedented strain on the healthcare system, emphasizing the need for rapid and effective COVID-19 treatment with complication management. Several investigations have found that a severe or critical progression of COVID-19 causes acute hypoxemic respiratory failure (AHRF), which necessitates a high fractional concentration of inspired oxygen (FiO_2_) and noninvasive ventilation (NIV) techniques such as a face mask, a non-rebreather mask (NRM) etc. [13-16]. HFNC, on the other hand, tends to be more successful than others because it can reach 100% humidification at 37°C and has a positive end-expiratory pressure (PEEP) effect while patients breathe with the mouth closed [17, 18].

The relation between COVID-19 and diabetes, as well as the condition’s long-term effects on people, is still being researched and investigated. This study focuses on seeing how HFNC with NRM compares to mechanical ventilation (MV) in diabetic COVID-19 patients hospitalized in different ICUs in Bangladesh. Our goal was to shed light on this technique’s usefulness in severe or critical instances where MV facilities are limited. The findings of this study can assist specialist doctors and the whole healthcare system of our country in expanding the range of treatment options available to individuals suffering from life-threatening COVID-19 consequences.

## 2. Materials and Methods

### 2.1. Sample size and data collection

A cross-sectional observational study was carried out with a sample of 240 COVID-19 patients who required medical attention in various medical facilities and were verified positive by Real-time Reverse-transcriptase Polymerase Chain Reaction (rRT-PCR) analysis.

Patients (diabetic or non-diabetic) with a requirement of HFNC with or without NRM were deemed candidates. As the primary sources of data, a pertinent questionnaire and medical history were used. All retrospective data gathered via telephone interviews were manually entered into an online format. All data entered on the questionnaire that matched the participants’ responses were double-checked before being posted and the recordings were stored.

### 2.2. Ethical approval

This research and its protocol were approved by the 250 Bedded Chattogram General Hospital’s Institutional Review Board (Approval No.: 1724).

### 2.3. Inclusion and exclusion criteria

COVID-19 subjects with 6.5% glycated haemoglobin (HbA1c) content, who had recently demonstrated any validated biochemical examination of diabetes mellitus were included in the diabetes cohort. Uncontrolled hyperglycemia was classified into two or more blood glucose level examinations that yielded a result more than 11.1 mmol/l, regardless of blood sugar levels.

Patients having dyspnea, i.e. a respiratory rate of ≥ 30 beats per minute in rest and sustained SpO_2_ less than 90% after receiving 15 liters per minute of oxygen were deemed candidates for HFNC. Besides, those who failed to maintain desired oxygen saturation (SpO_2_ >90%) after high flow were also given a face mask containing NRM and HFNC. They were enlisted in the ‘severe’ category. Those with respiratory failure, sepsis, or shock, which necessitated MV, as well as those with multiple organ failures requiring ICU support, were placed in the ‘critical’ category. Patients who required MV or NIV from the start of their ICU stay, as well as those who refused to participate in the research, were excluded. The Berlin definition was used to specify acute respiratory distress syndrome (ARDS), and the Sepsis-3 criteria were utilized to define shock [19, 20].

### 2.4. Study sites

The research was carried out in five hospitals: the 250 bedded Chattogram General Hospital, Chittagong Medical College Hospital, Chattagram Maa-Shishu O General Hospital, and Parkview Hospital. These hospitals have dedicated general as well as intensive care units for the treatment and management of COVID-19 patients.

The cases’ epidemiological and demographic data were obtained by assigned investigators from the patients’ treatment records and interviews with the accompanying personnel. The study took place between April 15, 2021, and June 14, 2021.

### 2.5. rRT-PCR test

Throat swabs, nasopharyngeal swabs, and bronchial aspirates were obtained from patients and placed in a collection tube with a viral transport medium before being sent to the research laboratories. The SARS-CoV-2 RNA extraction for COVID-19 was carried out in the Molecular Biology laboratory of the Microbiology department of Chittagong Medical College in accordance with WHO guidelines [21].

### 2.6. Statistical analysis

To check possible correlations between categorical variables, Pearson’s Chi-Square (χ2) (where <20% of cells had expected count less than 5) and Fisher’s Exact (where ≥20% of cells had expected count less than 5) evaluation methods were used. Categorical and continuous variables were tested for associations by applying Independent-Samples T-Test (95% confidence interval) and ‘means’ with ‘standard deviations’ were compared. *P* values less than 0.05 were considered statistically significant. *P* value of “Equal variance not assumed” was considered in case of categorical and continuous variable correlation. Factors that had significant differences when correlating with diabetic/non-diabetic group were further analyzed by dividing into two groups HFNC only and HFNC + NRM treated patients. Then the statistically significant factors were analyzed against HFNC only and HFNC + NRM by Simple bivariate logistic regression and multiple bivariate logistic regression to find the significant factors, crude odds ratio (COR), adjusted odds ratio (AOR), and their ranges at 95% confidence interval. Omnibus tests of model coefficients’ *P values* less than 0.05 and Hosmer and Lemeshow Goodness of Fit test’s *P values* greater than 0.05 were considered significant to test if the regression model had been fit for the data. Specificity and sensitivity of the data were also tested during regression analysis. All data analysis tests were performed in IBM SPSS version-25.

## 3. Results

### 3.1. Basic socio-demographic characteristics and Investigation result of the patients

***Table 1*** illustrates the basic demographic characteristics of HFNC treated patients. We found that the prevalence of diabetes among female patients (64.6%), those ≥ 50 years of age (64.1%), urban residents (57.9%), previously smokers (56.0%), and those who never smoked (57.6%) within the study sample (***Table 1***). Age was significantly related to diabetes mellitus (p< 0.001). In this data, 20.8% (50/240) patients did not have any comorbidities (p< 0.001). Among the hypertensive and IHD patients, 31.7% and 29.8% were non-diabetic, respectively (p< 0.001 & p= 0.037) (***Table 1***). Persistence of fever had a significant association with diabetes mellitus (p= 0.012), and 59.1% of the feverish patients had diabetes. Other than fever, cough (72.5%; 174/240) and breathlessness (67.5%; 162/240) were common symptoms. Among 240 patients who comprised the study sample, 47.1% (113) patients were given HFNC and NRM oxygenation simultaneously, and 52.9% (127) patients were treated with HFNC only (***Table 1***). Among the 126 patients who died during the study period, 56.3% were diabetic. For diabetic cases, the duration (in days) between the first onset of COVID-19 associated symptoms and death was higher (17.48 ± 7.15) (p= 0.006). Additionally, for diabetic patients the stay in the ICU was longer (9.06 ± 5.70) as compared to the non-diabetic patients (7.41 ± 5.11) (p= 0.020) (***Table 1***).

**Table 1:**
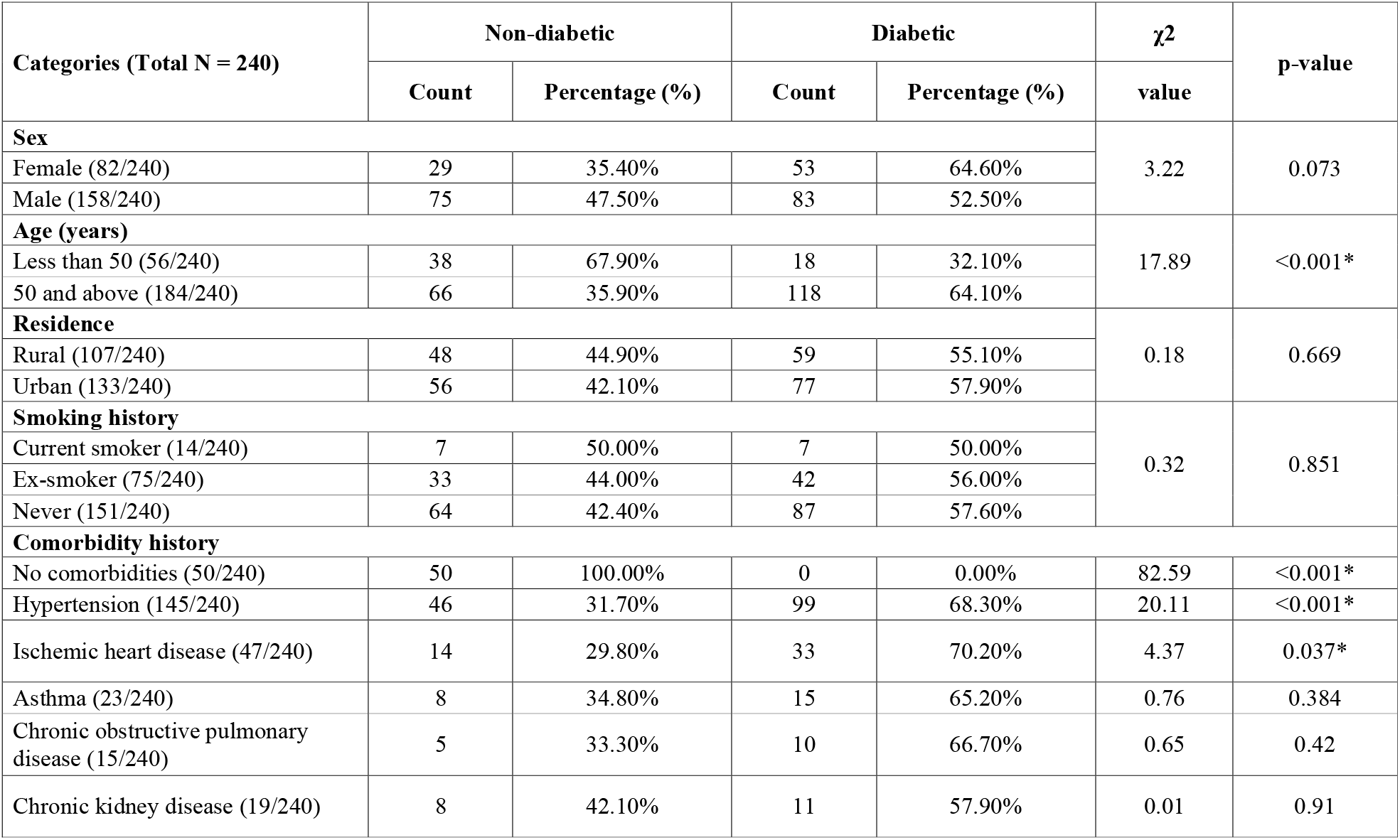
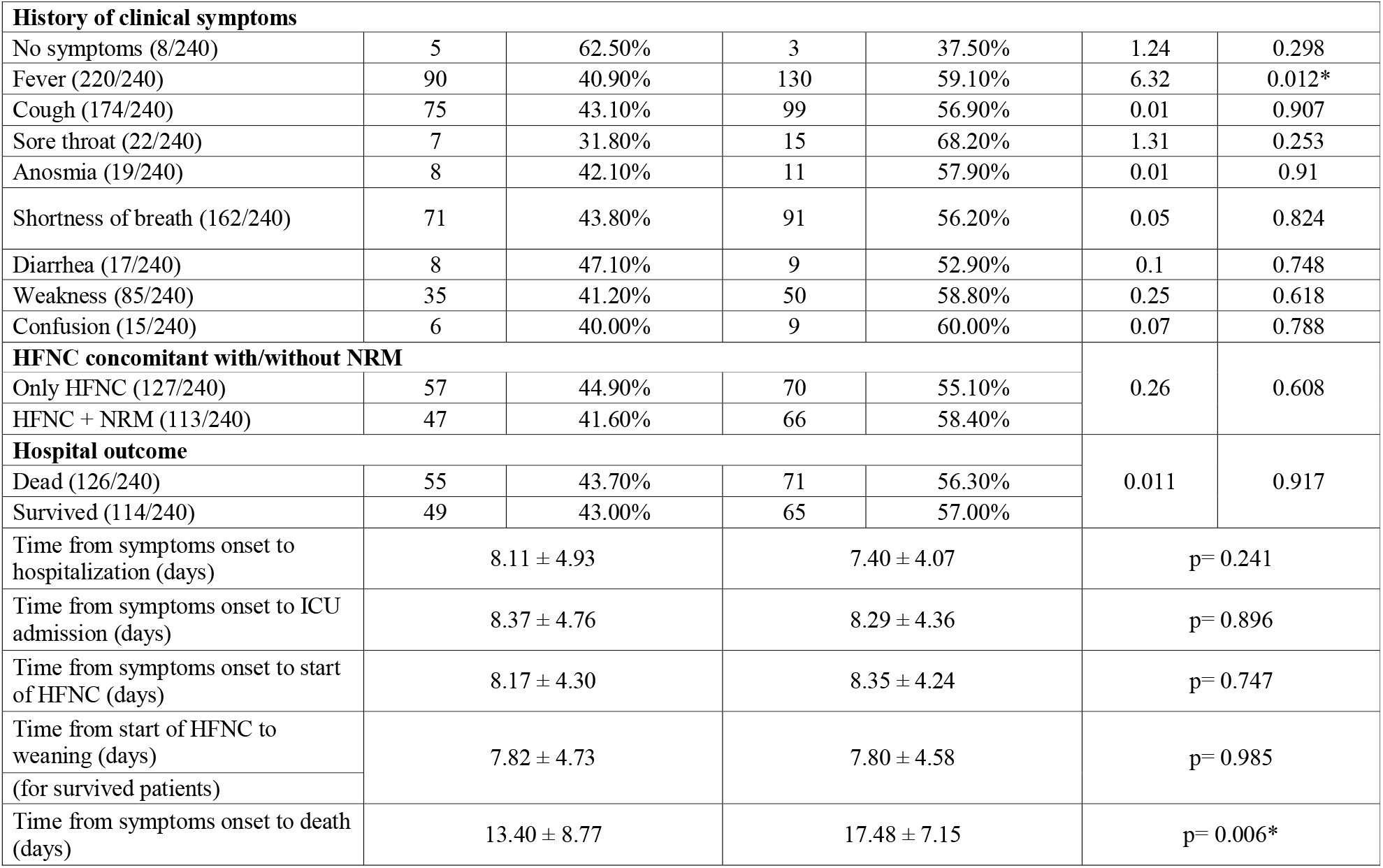
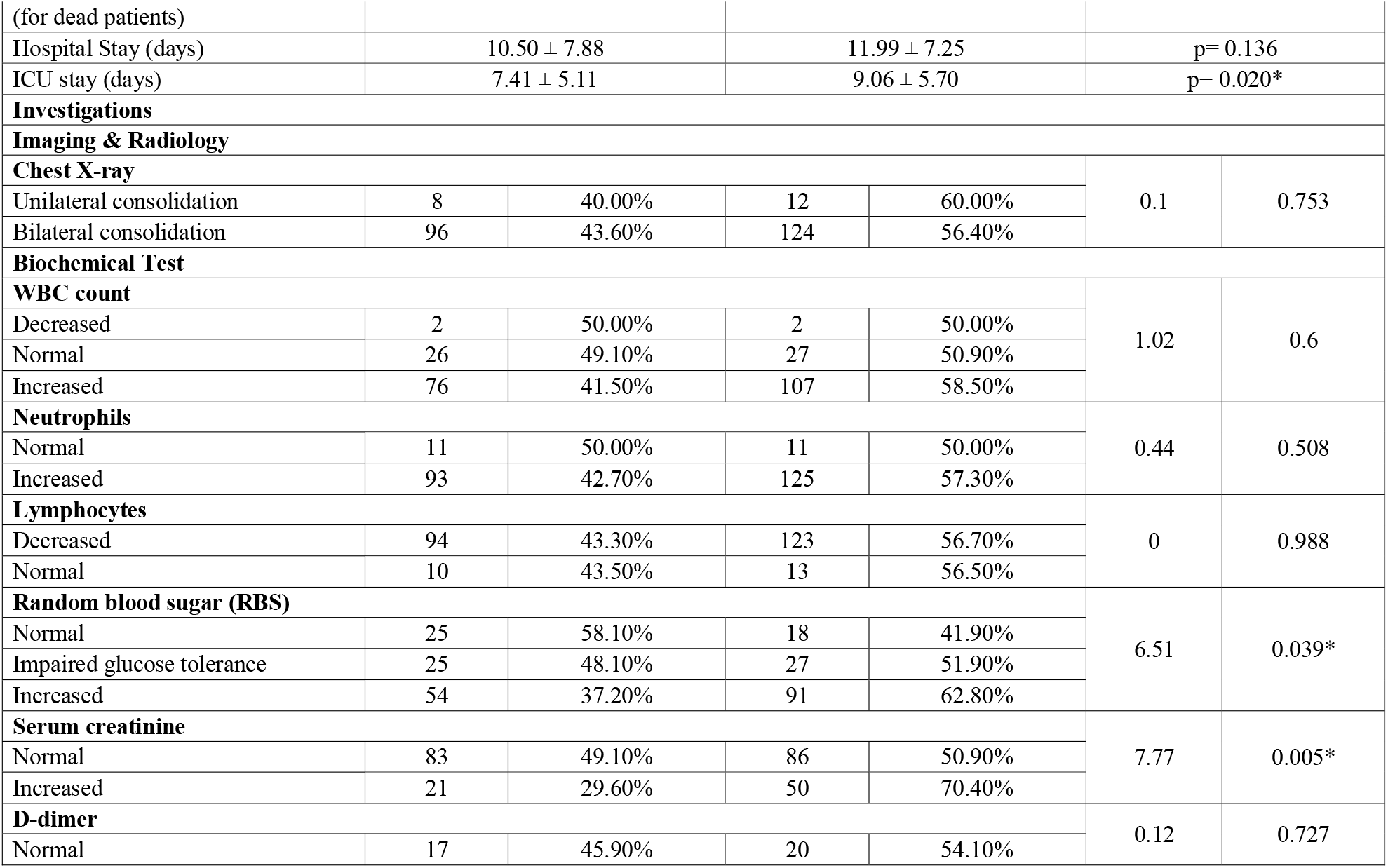
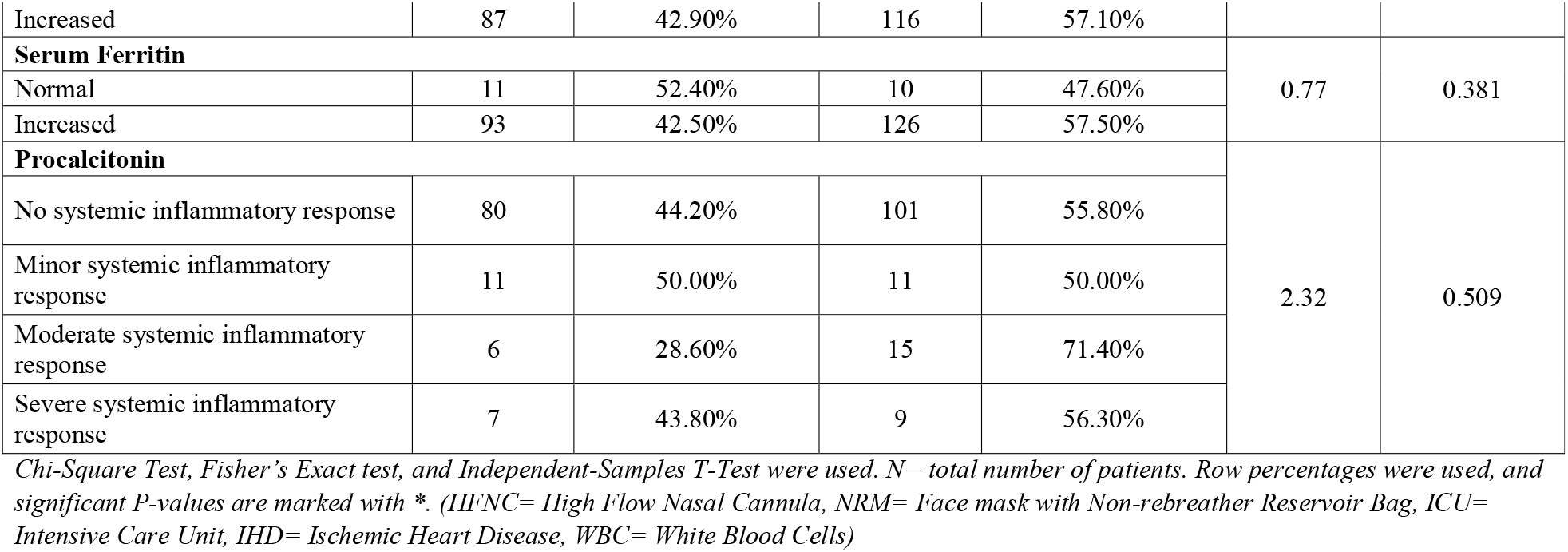
Basic Demographic Characteristics and Investigations of patients treated with HFNC.

Data obtained from each patient’s investigation report has been included in ***Table 1***. Random blood sugar (RBS) and serum creatinine levels were significantly related to diabetes mellitus (p= 0.039 & p= 0.005). Amongst those with high creatinine level, 70.4% were diabetic, and 29.6% were non-diabetic (***Table 1***).

### 3.2. Immediate complications of the patients

***Supplementary Figure 1*** (a) and (b) representing the complications of HFNC, show that non-visible nasal bleeding followed by nasal obstruction by clotted blood was the most observable complication. We found that in the study sample, non-diabetic patients (76/104; 73.1%) suffered from HFNC complications more than diabetic patients (96/136; 70.6%) (***Supplementary Figure 1***). In this study, 38.2% of the diabetic patients and 42.3% of the non-diabetic patients had the aforementioned complications before their death. Other complications of HFNC were headache (59/240; 24.6%), discomfort (71/240; 29.6%), and frequent displacement of the nasal cannula (89/240; 37.1%) (***Supplementary Figure 1***). In addition, complaints relating to the irritation in the nostrils were also reported by the patients, which is considered as an indication of discomfort in the current survey.

### 3.3. Impact of HFNC and other treatments

The impact of HFNC and NRM on ICU admitted COVID-19 induced AHRF patients is illustrated in ***Table 2***. The partial pressure of oxygen (mmHg) after the administration of HFNC (only) was significantly (p= 0.031) higher for the non-diabetic patients (69.30 ± 23.56) than those with diabetes (61.50 ± 14.49) (***Table 2***).

**Table 2:**
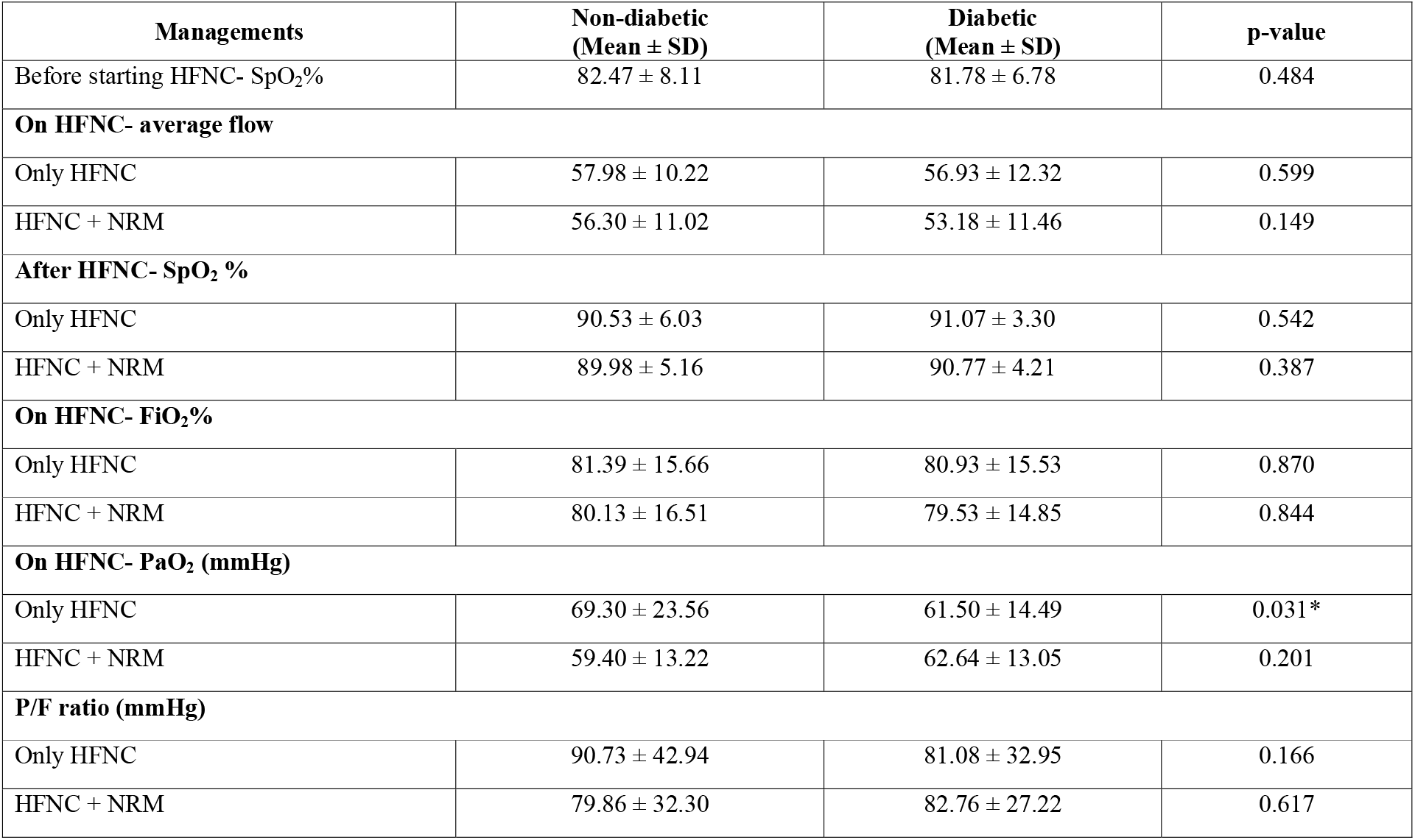
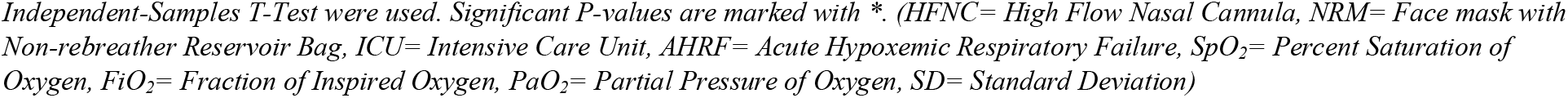
Impact of HFNC (with/without NRM) for ICU admitted COVID-19 patients.

***Table 3*** describes the impact of HFNC (with/without NRM) on diabetic and non-diabetic patients with COVID-19 induced AHRF. Among the patients who were treated with both HFNC and NRM, the prevalence of AHRF was higher for those aged ≥ 50 years with diabetes (67.4%) (p< 0.001). Elderly diabetic patients needed HFNC concomitant with NRM (***Table 3***). Most of the patients who did not have any comorbidity (62.0%; 31/50) were managed with HFNC only (p< 0.001). Higher proportions of diabetic patients having hypertension had to be treated with both HFNC (singularly) (70.1%) and HFNC combined with NRM (66.7%) than the non-diabetic hypertensive cases (***Table 3***). A total of 58.4% of the patients treated with only HFNC were diabetic and had elevated body temperature (p= 0.034). Besides, most of the patients who needed both HFNC and NRM had raised RBS (73.8%; p= 0.001) and creatinine levels (75.7%; p= 0.009) (***Table 3***).

**Table 3:**
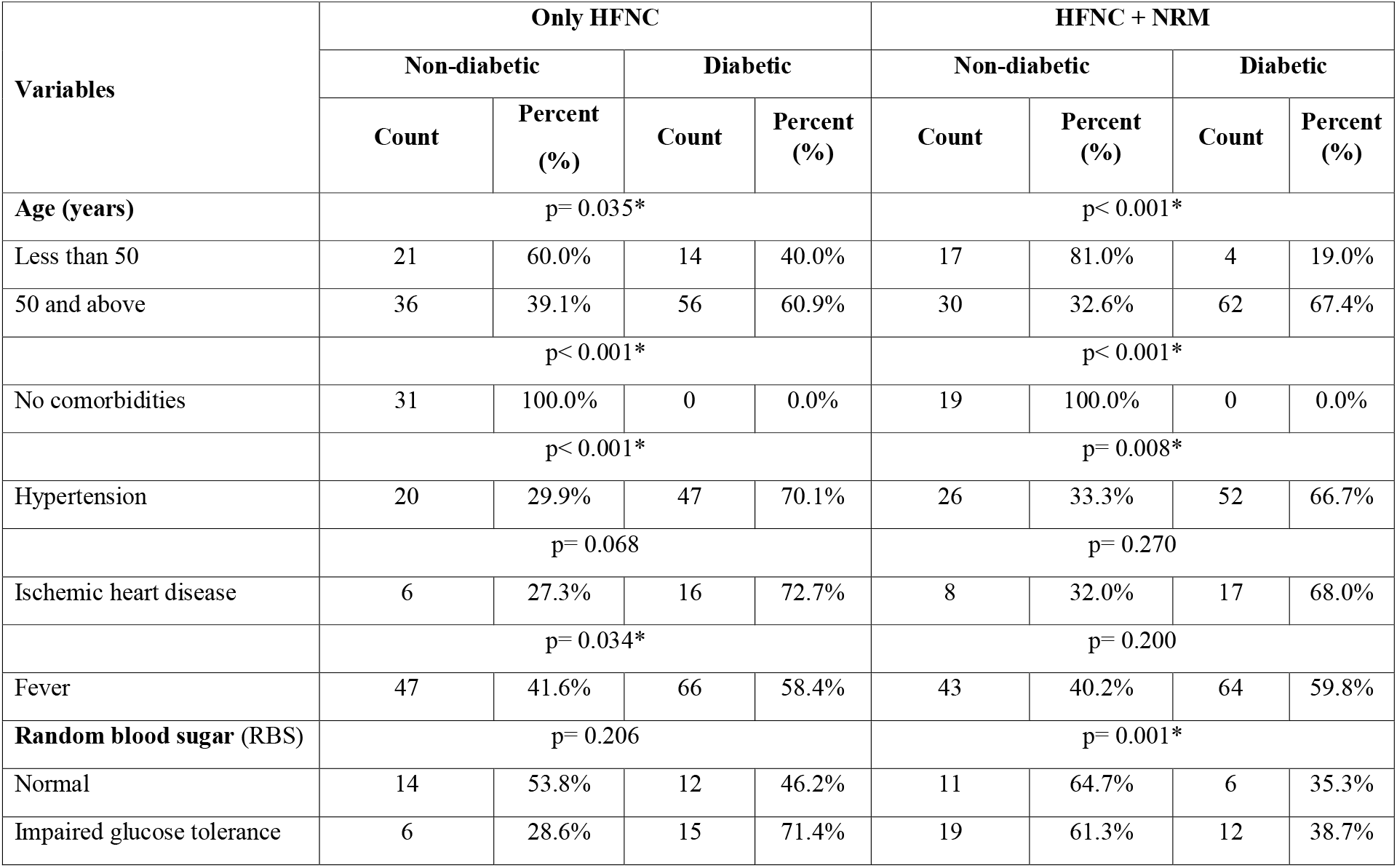
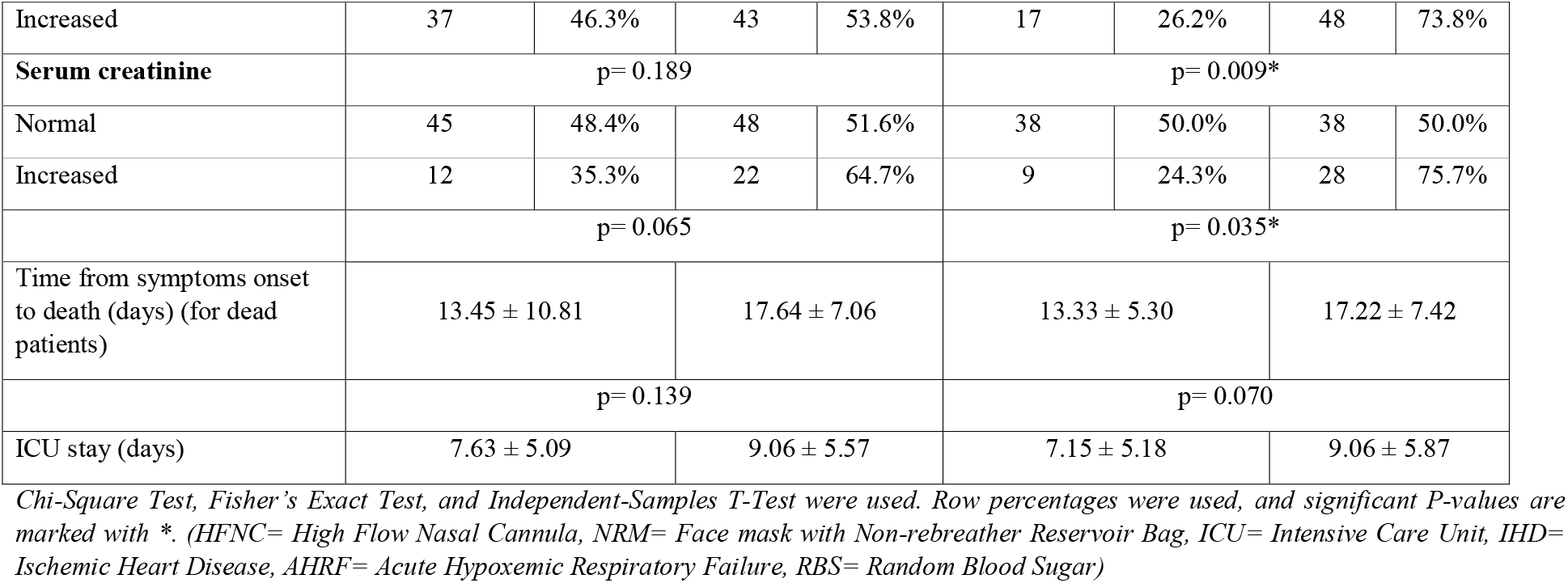
Impact of HFNC (with/without NRM) for ICU admitted COVID-19 (Diabetic/non-diabetic) patients.

***Figure 1*** (a) and (b) show the treatment protocols alongside HFNC (with/without NRM) for the diabetic and non-diabetic patients, respectively. The protocol included antivirals, antibiotics, steroids, low molecular weight heparin, interleukin-6 inhibitor (Tocilizumab), and convalescent plasma therapy (***Figure 1***). Among patients with diabetes, the survival ratio after only HFNC was higher for those who were given oral (100.0%) antiviral drugs than those administered in intravenous (IV) (35.3%) form. But the response rate of IV antivirals (58.7%) increased when NRM was also used for treating them. On the contrary, the survival rate was high after using IV antivirals beside HFNC concomitant with (50.0%)/without (75.0%) NRM amongst the patients not having diabetes (***Figure 1***). The response rate of IV antibiotics to survival/death was not significantly different after HFNC administration with/without NRM for patients not/having diabetes. When plasma therapy was given with both HFNC and NRM, the survival rate was significantly high among non-diabetic patients with COVID-19 induced AHRF (100.0%). Another observation was a high survival rate among the diabetic patients after giving dexamethasone with HFNC + NRM (62.7%) (***Figure 1***). The majority of non-diabetic patients (62.5%) having administered both HFNC and NRM survived when they got tocilizumab treatment. Though the survival percentages among non-diabetic patients after HFNC (with/without NRM) and heparin were almost similar, a twice-daily dose of heparin with HFNC + NRM could save 63.6% of the diabetic patients (***Figure 1***).

**Figure 1:**
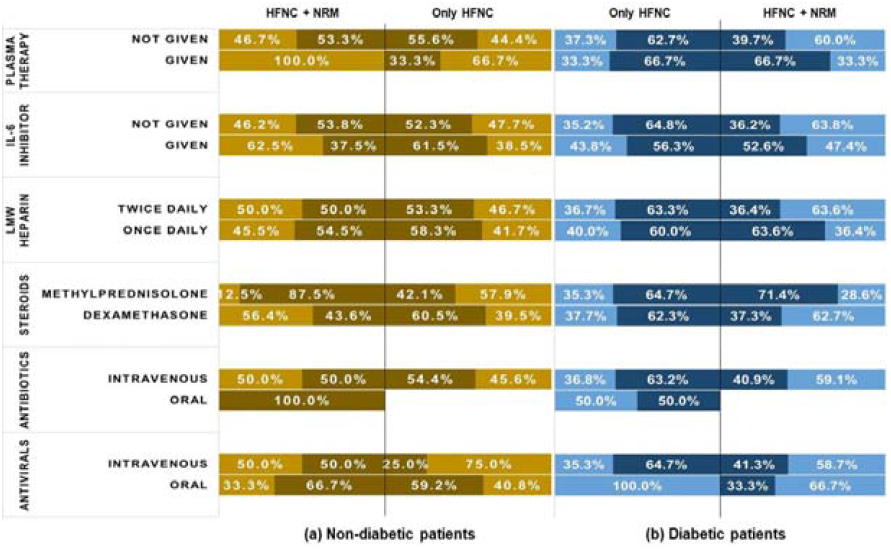
Brown (a) and blue (b) represent the treatments for non-diabetic and diabetic patients, respectively. Light and dark shades of (a) and (b) indicate survival and death rate after the treatments. One side of the black divider of each color denotes the percentage of patients treated with HFNC only, while another side signifies HFNC + NRM treated patients’ percentage. HFNC= High Flow Nasal Cannula, NRM= Non-Rebreather Mask, IL-6 inhibitor= Interleukin-6 inhibitor, LMW Heparin= Low Molecular Weight Heparin

### 3.4. Factors associated with HFNC with/without NRM

Non-diabetic patients who were managed with only HFNC were 6.5 (1.3-33.1) times less feverish than diabetic patients. The chance of having IGT was about twelve times (AOR= 12, 1.1-129.8) high for the diabetic COVID-19 patients who were also given only HFNC to maintain their oxygenation (***Table 4***).

**Table 4:** Factors associated with HFNC only and HFNC+NRM management of ICU admitted diabetic COVID-19 patients.

| Variables | Only HFNC |  | HFNC + NRM |  |
| --- | --- | --- | --- | --- |
|  | COR with range (95% CI) | AOR with range (95% CI) | COR with range (95% CI) | AOR with range (95% CI) |
| Age (in years) |  |  |  |  |
| Less than 50 (ref) | 1.0 | 1.0 | 1.0 | 1.0 |
| 50 and above | **2.3 (1.1-5.2)\*** | 2.5 (0.8-8.5) | **8.8 (2.7-28.4)\*** | **5.8 (1.1-31.2)\*** |
| Comorbidity history |  |  |  |  |
| No comorbidities | 0.0 (0.0) | 0.0 (0.0) | 0.0 (0.0) | 0.0 (0.0) |
| Hypertension | **3.8 (1.8-7.9)** | 0.7 (0.2-2.2) | **3.0 (1.3-6.8)\*** | **0.1 (0.01-0.8)\*** |
| Clinical sign |  |  |  |  |
| Fever | **3.5 (1.0-11.9)\*** | **6.5 (1.3-33.1)\*** | 3.0 (0.5-17.0) | 2.2 (0.3-18.7) |
| Random blood sugar |  |  |  |  |
| Normal (ref) | 1.0 | 1.0 | 1.0 | 1.0 |
| Impaired glucose tolerance | 2.9 (0.9-9.9) | **11.7 (1.1-129.8)\*** | 1.2 (0.3-4.0) | 1.2 (0.3-5.9) |
| Increased | 1.4 (0.6-3.3) | 1.5 (0.4-5.0) | **5.2 (1.7-16.2)\*** | **5.1 (1.2-20.8)\*** |
| Serum creatinine |  |  |  |  |
| Normal (ref) | 1.0 | 1.0 | 1.0 | 1.0 |
| Increased | 0.6 (0.3-1.3) | 0.6 (0.2-1.8) | **3.1 (1.3-7.5)\*** | 2.1 (0.7-6.3) |

*Simple and Multiple Bivariate logistic regression was used and p-values less 0.05 (marked with \* and are bold) are considered significant. (COR= Crude Odds Ratio, AOR= Adjusted Odds Ratio.)*
*\*\*Times from symptoms onset to death is removed from the analysis as it is strongly correlated with other factors.*

The chance of being aged at least 50 years was almost six (AOR= 6.2, 1.1-31.2) times higher among diabetic HFNC + NRM treated patients. Non-diabetic patients (HFNC + NRM treated) were more likely to have hypertension than diabetic ones. Moreover, among HFNC + NRM treated patients, diabetic patients were five times (AOR=5.1, 1.2-20.8) more likely to have increased blood glucose levels (***Table 4***).

## 4. Discussion

Because a previous study recommended the usage of HFNC for minimizing invasive/mechanical ventilation use [22], in this study, the clinical effect of HFNC as a mode of providing supplemental oxygenation to COVID-19 diabetic patients was observed to analyze whether the use of this mechanism is efficient enough to be reiterated on a large scale to reduce the burden of MV support in the context of Bangladesh’s COVID-19 landscape. When HFNC failed to maintain the optimum oxygenation with at least 92% of SpO_2_, NRM was also added to the ICU admitted patients. We also tried to find the success rate of using NRM concomitant with HFNC to the COVID-19 induced AHRF diabetic patients.

In this data, 136 among 240 HFNC treated AHRF patients had diabetes as comorbidity, which aligns with the statement by a study in China, which stated that diabetes mellitus is a commonly observed comorbidity in severe COVID-19 cases [23]. Diabetes among elderly patients was considered a risk factor for the severe prognosis of COVID-19 [11, 24]. Similar to a study in Bangladesh, the proportion of diabetic COVID-19 patients was significantly higher among those aged ≥ 50 years [12]. The findings of this survey showed that a great proportion (almost 6 times) of the elderly diabetic patients needed both HFNC and NRM to maintain their oxygenation because they could not maintain the optimum oxygen level with HFNC only.

Analogous to previous studies, we found a higher mortality rate among COVID-19 patients with type-2 diabetes compared with the non-diabetic patients, establishing diabetes as a risk factor for increased mortality [23, 25, 26]. As per the current study’s findings, face masks with non-rebreather reservoir bags were given together with HFNC to 113 patients, and among them, 58.4% were diabetic. After using nasal cannula only, non-diabetic patients showed more improvement of PaO_2_ than the diabetic ones. So, to maintain oxygenation of the severely/critically ill COVID-19 diabetic patients, NRM was also needed along with HFNC.

Saha et al. asserted the high prevalence of hypertension among the diabetic COVID-19 patients, and it was an important factor in the progression of COVID-19 for severe/critical patients [12]. In this study, it was observed that hypertensive diabetic COVID-19 patients required NRM along with HFNC to maintain the oxygen saturation. Besides, a significant difference in the presence of heart disease among diabetic and non-diabetic patients was also found in this current study. High prevalence of fever was found among the AHRF patients with diabetes in this study and only HFNC treated diabetic patients being 6.5 times more feverish might prove that fever was common among diabetic patients. As all the patients in this study were admitted to the ICU due to hypoxemic respiratory failure following COVID-19, the duration of ICU stay of the diabetic and non-diabetic patients was also observed. This study found a noticeable difference between them. Diabetic patients had to stay in the ICU for a longer period than those who did not have diabetes.

Increased blood glucose level is established as a determinant in the pathogenesis of the infectious disease, like the SARS-CoV-2 virus, and can make the diabetic patients immunocompromised, leading to their critical conditions after SARS-CoV-2 infection [27, 28]. This data supporting these studies proved that most of the severely/critically ill patients faced a rise in blood sugar, and among them, more than 60% were previously diabetic. Moreover, among diabetic patients only HFNC treated ones showed a high odds ratio of having IGT and HFNC + NRM treated ones showed of having increased blood glucose level. This data might prove that those who had increased blood glucose level rather of having IGT needed both HFNC and NRM to maintain their oxygen level.

In a study in China, it was stated that COVID-19 patients gradually develop kidney dysfunctions/acute kidney injury (AKI) as SARS-CoV-2 uses ACE2 (angiotensin-converting enzyme II) as a cell entry receptor [29]. A PubMed database indicates that ACE2 RNA expressions in gastrointestinal organs (small intestine, duodenum) and urinary organs (kidney) are much higher (nearly 100-fold) than that in lungs [30]. Compliant with these data, this current study noticed that increased serum creatinine level was found significantly among critically ill COVID-19 patients, and most of them did not have a previous history of kidney disease. Moreover, creatinine rise was high for those who were diabetic, and this indicates that elevation of creatinine level might be a determining factor of severity during SARS-CoV-2 infection for diabetic patients. As the severity of the disease for ICU admitted COVID-19 patients might increase because of diabetes and lately developed kidney dysfunction, HFNC and NRM both were needed to support most of the patients with raised sugar and creatinine levels.

Non-visible nasal bleeding along with mucosal obstruction was common for both diabetic and non-diabetic patients. So, to prevent this, liquid paraffin and normal saline were used. As this was a locally practiced procedure, more research is needed to establish the process for averting these complications. In a study, it was established that glucocorticoids can induce varying degrees of diabetes and this was similar to our data when patients were given steroids [31]. So, insulin was administered to all who were previously diabetic/ had been taking oral hypoglycemic drugs or insulin (switched from oral to injectable form)/ developed diabetes as a side effect of steroids. Dexamethasone with HFNC and NRM showed a good survival ratio for the diabetic patients, but convalescent plasma therapy worked effectively (with HFNC and NRM) for the non-diabetic patients showing a survival rate of 100.0%.

## 5. Conclusion

This study was conducted for analyzing the clinical outcomes of HFNC with/without a NRM on severely ill COVID-19 diabetic patients. As per the findings, the majority of the elderly diabetic and hypertensive diabetic patients needed both HFNC and NRM to sustain their oxygenation. Furthermore, increased blood sugar might prove that it may be a determining factor for the need of HFN + NRM for COVID-19 induced AHRF diabetic patients. As it is a multicentric prospective study, the findings of this study are representative of the situation of most hospitals in Bangladesh. As HFNC with/without NRM as per this study, was found to have association with a significant clinical improvement in severe case of COVID-19 in both diabetic and non-diabetic cases, the burden on MV and clinical demand of MV in constrained clinical settings can be to some extent reduced by considering HFNC with/without NRM and other therapeutics as an efficient candidate for supplemental oxygenation.

## Data Availability

All data produced in the present study are available upon reasonable request to the authors

## Abbreviation

HFNC: High Flow Nasal Cannula
NRM: Face Mask with Non-Rebreathing Reservoir Bag/Non-Rebreather Mask
COVID-19: Coronavirus Disease 2019
ICU: Intensive Care Unit
RBS: Random Blood Sugar
SARS-CoV-2: Severe Acute Respiratory Syndrome related Coronavirus
AHRF: Acute Hypoxemic Respiratory Failure
FiO_2_: Fractional Concentration of Inspired Oxygen
NIV: Non-Invasive Ventilation
MV: Mechanical ventilation
SpO_2_: Percent Saturation of Oxygen
IHD: Ischemic Heart Disease
rRT-PCR: Real Time Reverse Transcriptase-Polymerase Chain Reaction
WHO: World Health Organization
WBC: White Blood Cell
PaO_2_: Partial Pressure of Oxygen
SD: Standard Deviation

## Declarations

### Authors’ Contributions

AFM Tareq Bhuiyan: Design, clinical studies, data acquisition; Sudipta Deb Nath: Data acquisition, data analysis, manuscript preparation; Md Jakir Hossain: Concepts, data analysis, manuscript review; Shuva Das: Experimental studies, manuscript preparation; Moumita Das: Concepts, clinical studies; Moinul Ahsan: Statistical analysis; Md. Iftekher-E-Alam Ziad: Clinical studies; Fahmida Khatun Padma: Clinical studies; Rana Dey: Clinical studies; AKM Shamsul Alam: Manuscript editing; Farial Hoque Zehan: Clinical studies; Ayan Saha: Design, Manuscript preparation

## Acknowledgements

Authors would like to thank the authorities of 250 bedded Chattogram General Hospital, Chittagong Medical College Hospital, Chattagram Maa-Shishu O General Hospital, Parkview Hospital, and Surgiscope Hospital Ltd.

---

**Supplementary Figure 1:Immediate complications of (a) diabetic and (b) non-diabetic patients, accordingly.**

X and Y axis indicate different complications and % of patients, respectively. Blue and red shades individually signify death and survival rates of the patients having faced the particular complication. N= total number of patients suffered from the specific complication.

