## Supplementary figures and images for "Clinical observation of high-flow nasal cannula with non-rebreather mask use on severe or critically ill COVID-19 diabetic patients"

### Supplementary Figure 1

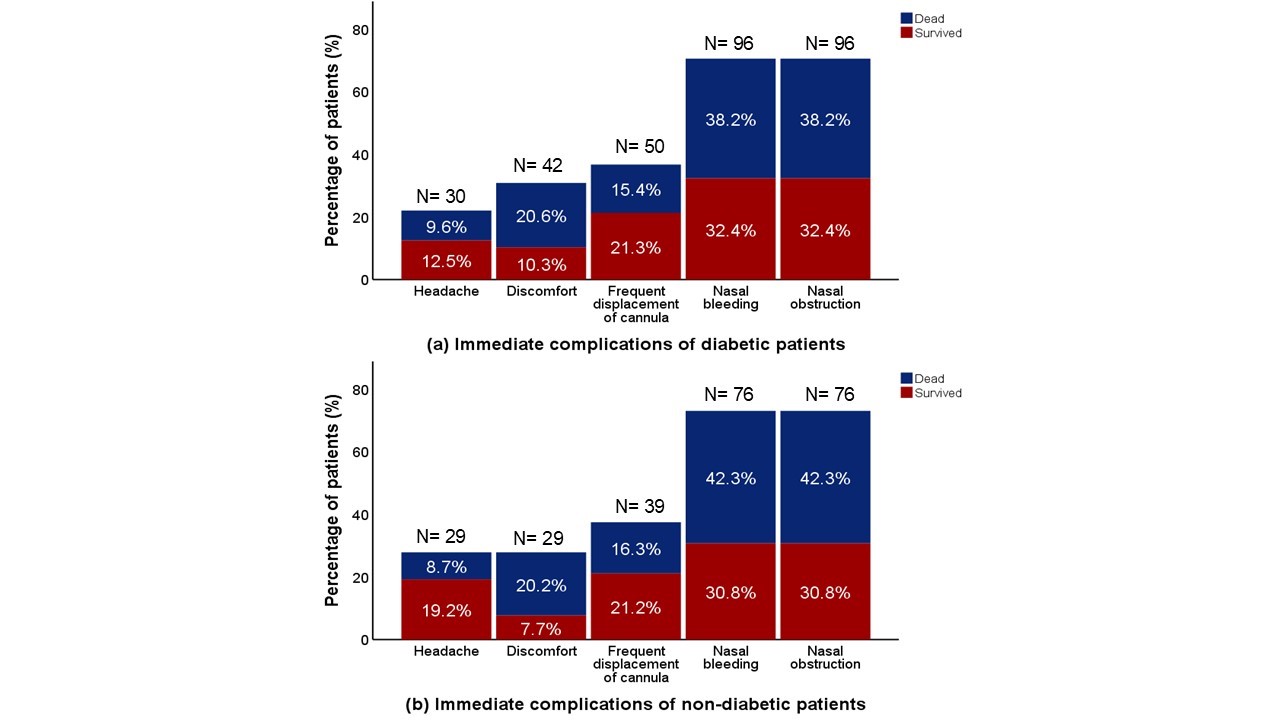
